# Less invasive SARS-CoV-2 testing for children: A comparison of saliva and a novel Anterior Nasal Swab

**DOI:** 10.1101/2022.09.21.22280208

**Authors:** Shidan Tosif, Lai-yang Lee, Jill Nguyen, Chris Selman, Anneke C Grobler, Alissa McMinn, Andrew Steer, Andrew Daley, Nigel Crawford

## Abstract

Reducing procedural discomfort for children requiring respiratory testing for SARS-CoV-2 is important in supporting testing strategies for case identification. Alternative sampling methods to nose and throat swabs, which can be self-collected, may reduce laboratory-based testing requirements and provide rapid results for clearance to attend school or hospital settings. The aim of this study was to compare preference and diagnostic sensitivity of a novel anterior nasal swab (ANS), and saliva, with a standard combined nose and throat (CTN) swab. The three samples were self-collected by children aged 5-18 years who had COVID-19 or were a household close contact. Samples were analysed by reverse transcription polymerase chain reaction (RT-PCR) on the Allplex SARS-CoV-2 Assay. Most children and parents preferred the ANS and saliva swab over the CTN swab for future testing. The ANS was highly sensitive (sensitivity 1.000 (95% Confidence Interval (CI) 0.920, 1.000)) for SARS-CoV-2 detection, compared to saliva (sensitivity 0.886, 95% CI 0.754, 0.962). We conclude the novel ANS is a highly sensitive and more comfortable method for SARS-CoV-2 detection when compared to CTN swab.

## Introduction

Self-collection is a preferred method of respiratory sample collection for SARS-CoV-2 detection^1^. Collecting samples in children presents challenges due to discomfort and poor compliance that may impact accuracy and parental confidence in testing^2^. Less invasive samples such as saliva can be taken, however studies reflect lower sensitivity in SARS-CoV-2 detection compared with nasal swabs^3,4^. Anterior nasal swabs are acceptable alternatives to nasopharyngeal swabs^5^, and a novel flocked anterior nasal swab (ANS) with features to reduce discomfort has been designed for children^6^. In this study, we compared self-collected combined nose and throat (CTN), saliva and novel anterior nasal swabs for detection of SARS-CoV-2 by polymerase chain reaction (PCR) and investigated feasibility in children aged 5-18 years.

## Methods

### Study design

The study was conducted at the Royal Children’s Hospital Melbourne (RCH), between February and May 2022. Self-collected CTN, ANS and saliva swabs were obtained concurrently. Swabs were collected by the patient or assisted by the parent, without clinician assistance. Swabs were collected at home, Emergency Department, or inpatient ward. Informed consent was obtained prior to swab collection. Ethics approval was by The Royal Children’s Hospital Human Research Ethics Committee (HREC77305). This trial was registered on ClinicalTrials.gov (NCT05043623).

### Participants

Children between the ages of 5-18 years with confirmed COVID-19 in the prior 7 days, or household contacts of these confirmed cases, were invited to participate.

### Test Methods

Parents/guardians followed written instructions for collection of the samples. The CTN samples were collected by swabbing the tonsillar beds and back of the throat for 3-5 seconds, followed by bilateral nasal insertion and rotation 5 times against the nasal wall with a flocked swab (FLOQSwab 551C, Copan, Brescia, Italy). Saliva samples were obtained by sucking a flocked swab for 30 seconds (FLOQSwab 552C, Copan, Brescia, Italy). The ANS was collected by inserting and leaving a Rhinoswab Junior (Rhinomed, Melbourne, Australia) in the anterior nares for 60 seconds, followed by side-to-side movements for 15 seconds. The order in which the swabs were taken was at the discretion of the child and parent/guardian.

### Acceptability Evaluation

Acceptability was assessed by an electronic survey following the swabs. A 5-point Likert scale or Wong-Baker FACES scale^7^ were used to rate comfort by the child (self-report) or parent.

### Analysis

In the laboratory, each sample was eluted into 500ul of phosphate buffered saline (PBS). The saliva and CTN samples were eluted through swirling. Due to the shape of the ANS, these samples could not be swirled, and instead eluted through vortexing and a pulse spin. All samples were extracted on Roche MagNA Pure 96 system using MagNA Pure 96 DNA and Viral NA Small Volume Kit. Samples were tested on the Allplex SARS-CoV-2 Assay (Seegene, Seoul, South Korea) which detects 4 target genes for SARS-CoV-2 (E, RdRP/S, N). Cycle thresholds (Ct) values for E-gene were recorded, and results were reported as “detected”, “not detected” or “inconclusive” in accordance with RCH laboratory standard operating procedure. E-gene was chosen of the four targets based on previous research for its use as a first-line target^8^.

### Statistical analysis

Data was recorded in REDCap^9,10^ before analysis in Stata (Version 17.0)^11^.The sensitivity of ANS and saliva samples were calculated and reported with 95% confidence intervals (CIs). When no virus was detected in one sample, but was present in the other, the Ct value was set at the maximum number of cycles performed in the lab for the undetected sample (38.73). The median difference in Ct values between the ANS and the standard CTN swab was compared. A boxplot was used to compare the distribution of the Ct values using the E gene among all sampling methods.

## Results

53 participants were recruited and eligible for the study. One participant did not complete the survey. Median age was 10.7 years (IQR 7.8-14.3) and tested 3.0 (IQR 1.0-5.0) days following COVID-19 diagnosis or exposure. At time of recruitment, 43 (82.7%) were confirmed SARS-CoV-2 positive and 9 were (17.3%) household contacts. 11 (20.8%) participants were asymptomatic. There were 44/52 PCR SARS-CoV-2 detections from the CTN (Table 1). All participants who tested positive to SARS-CoV-2 on the CTN were also detected by the ANS (sensitivity 1.000 (95% CI 0.920, 1.000)). The ANS identified one additional SARS-CoV-2 detection not detected by CTN. Of the saliva samples, 39/44 detected SARS-CoV-2 when compared to CTN (sensitivity 0.886, 95% CI 0.754, 0.962). ANS samples had a median Ct value difference of -1.4 (95% CI -2.4, -0.5), compared to 7.1 (95% CI 5.6, 9.4) for saliva, compared with CTN (Figure 1). Comfort scores for ANS and saliva were consistently “comfortable” to “a little uncomfortable” compared with CTN scores of “more” to “really” uncomfortable (see Figure 2). Amongst the three methods, saliva was the preferred sample for half of children 26/51 (51.0%), the ANS 21/51 (42.9%) and CTN 0/51 (0%). Between ANS and CTN for future testing, ANS was preferred by children 44/50 (88.0%) and parents 37/50 (74.0%).

**Figure 1:**
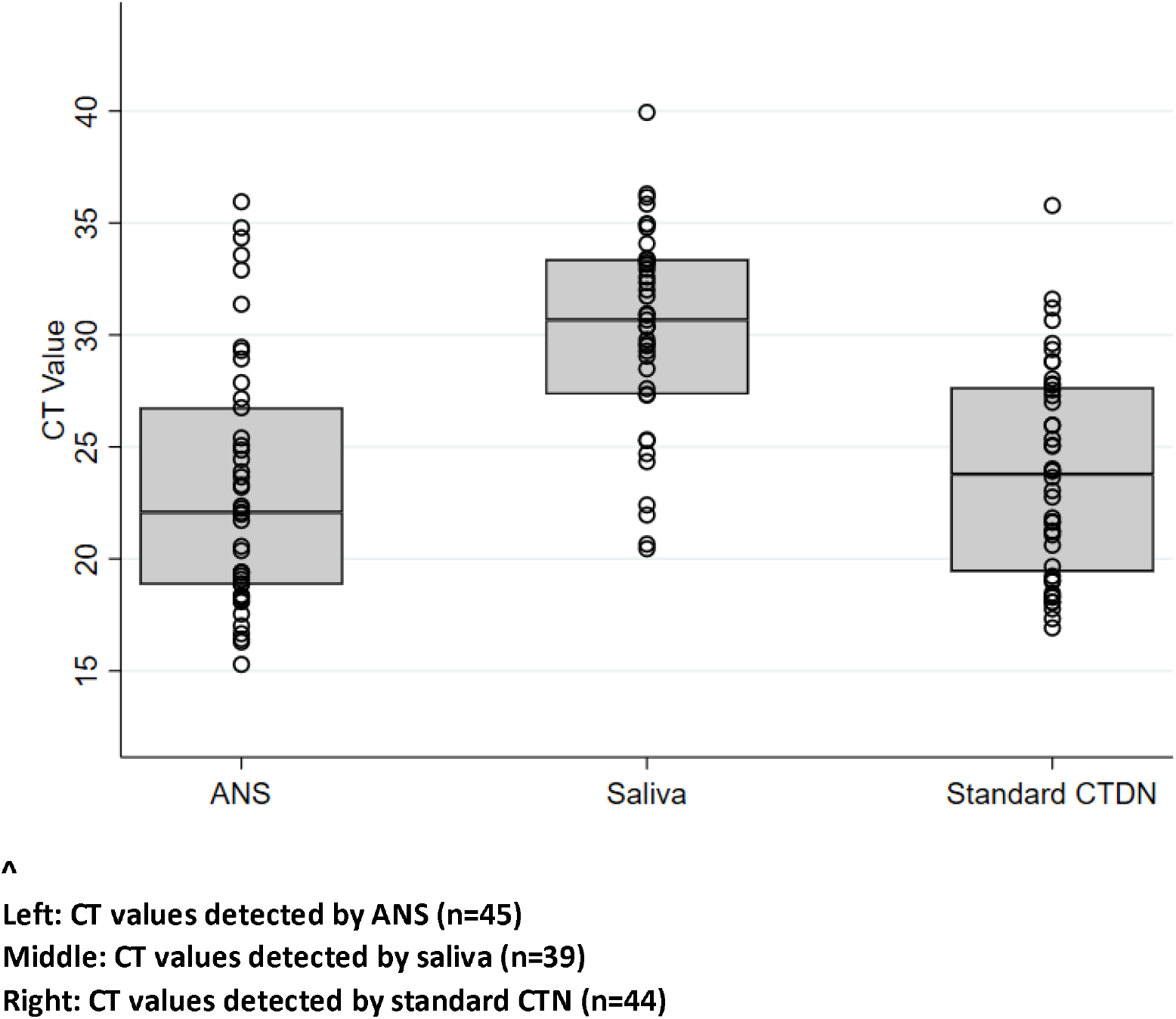
Distribution of CT-values for E-gene among all sampling methods.

**Figure 2:**
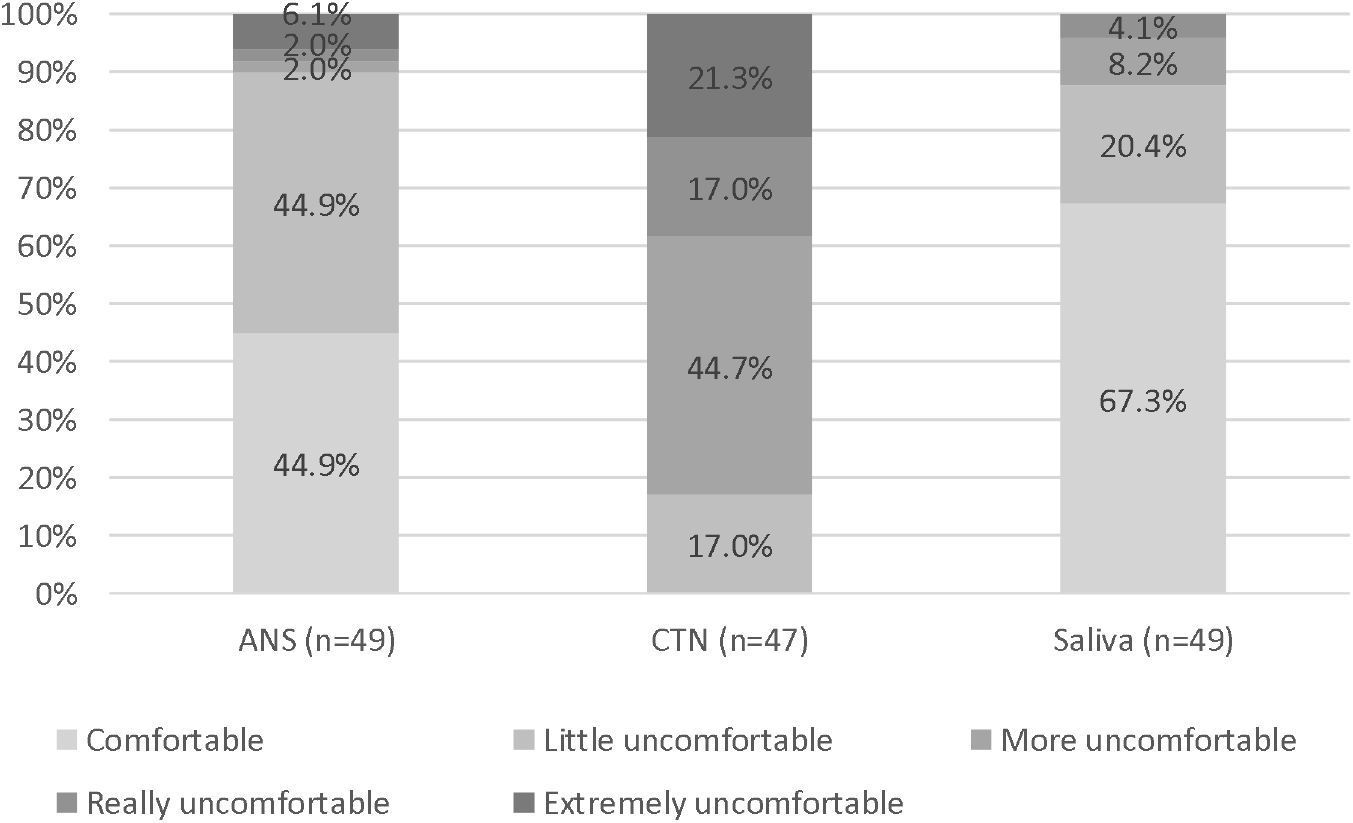
Comfort Rating of ANS, CTN and Saliva.

**Table 1.** Sensitivity of sampling methods using standard CTN as the reference comparator

| Sample | Result on ANS/Saliva in those detected on standard CTN |  | Sensitivity (95% CI) |
| --- | --- | --- | --- |
|  | Detected | Not detected |  |
| ANS | 44 | 0 | 1.000 (0.920, 1.000) |
| Saliva | 39 | 5 | 0.886 (0.754, 0.962) |

## Discussion

The novel ANS demonstrated high sensitivity with CTN for SARS-CoV-2 detection when compared with saliva. The saliva and ANS had similar comfort and preference scores and were both preferred over the CTN swab.

Self-collected respiratory samples for SARS-CoV-2 improve access and reduce risk of onward exposure to others^12-16^. However, standardisation of anatomical site of collection, adequate sample collection and time taken for sample delivery to the laboratory are common challenges^17^. Additional considerations are required for children undertaking respiratory testing as discomfort and procedural anxiety may impact testing success. In this study, the high sensitivity of the novel ANS combined with a favourable comfort profile are strengths of this method of collection. These factors are particularly important in children who may need frequent testing in school or hospital settings. However, adjustment by the laboratory was required for the novel ANS as it is shaped differently from the typical flocked swab and therefore the technique used for sample elution deviated from routine practice.

Saliva has been described as the patient preferred sample for SARS-CoV-2 detection compared with CTN, despite lower sensitivity^18-20^. However, there are different methods of saliva collection, including sucking on a collection device, oral swab or drooling into a container. Depending on how the sample is collected, processing of the samples in the laboratory may need to be adapted^21^. Collection of saliva by drool is challenging in younger children due to compliance. The sucked saliva swabs used in this study were an easy sample to collect and were more comfortable. This method is a feasible option for some children, such as those with special needs, where other methods may not be tolerated, and a lower sensitivity threshold is accepted.

Limitations of the study include adherence to written instructions was not monitored, thus there may be differences in collection technique, although this reflects real life testing. The order of the swabs was at the patient’s discretion, and this could affect the sample quality. Further research is needed to validate saliva and ANS for other respiratory viruses, and other home-based testing platforms such as rapid antigen testing.

## Conclusion

The novel ANS provides a more sensitive method for detection of SARS-CoV-2 in children than saliva with similar comfort scores by children and parents. Further research investigating methods of respiratory viral testing which reduce discomfort in children whilst providing accurate diagnosis of respiratory viruses are needed.

## Data Availability

All data produced in the present study are available upon reasonable request to the authors

## Abbreviations

ANS: Anterior Nasal Swab
COVID-19: Coronavirus disease 2019
CTN: Combined Throat and Nose Swab
RT-PCR: Reverse transcription polymerase chain reaction
SARS-CoV-2: severe acute respiratory syndrome coronavirus 2

## Acknowledgements

We thank the study participants and families for their involvement in this study. We also acknowledge the SAEFVIC Research Team (Hayley Giuliano, Chelsea Bartel and Anna Wylie). ST is supported by a Murdoch Children’s Research Institute Clinician Scientist Fellowship.

